# Sedentary Habits and Obesity: The Role of TV, Unemployment, and White-Collar Work on Obesity in Reproductive-Aged Women in Bangladesh

**DOI:** 10.1101/2024.09.24.24314338

**Authors:** Md. Zakiul Alam, Isna Haque Sheoti

## Abstract

**Introduction:** In Bangladesh, overweight and obesity are increasingly alarming, especially among women. Thus, this paper aims to explore whether television watching, white-collar jobs, and unemployment are working as sedentary behavior to increase overweight and obesity in Bangladesh.

**Data and methods:** We utilized cross-sectional data from the Bangladesh Demographic and Health Survey, 2017-2018. The dependent variable of the study was the nutritional status of women using three different measures: body mass index (BMI), Asian body mass index (ABMI), and tri-ponderal mass index (TMI). The frequency of watching television (TV) and types of employment were the two predictors of overweight and obesity in this study. The Multilevel Ordered Logistic Regression Analysis was used in this study.

**Results:** Women who have watched television at least once a week are more likely to have a BMI ≥25 and become overweight (30.3%) and obese (9.3%) than those who did not watch TV at all (20.1% and 3%). This same pattern of BMI is discernible among unemployed women (28.7% and 8.2%) compared to women in agricultural work (16% and 2.9%). Professional women have the highest risk of being overweight and obese (35.4% and 10.9%). The pattern of overweight and obese is consistent for ABMI and PMI. However, these indicators (ABMI & PMI) report a higher prevalence of overweight and obesity than BMI.

**Conclusion:** In this era of technology, TV watching, and other technology will increase with time. More and more women are engaging in white-collar jobs or looking for jobs. In these circumstances, policymakers need to focus on how this use of technology can decrease overweight and obesity among women. More studies need to be conducted to explore the effect of other sedentary behaviors and take steps based on those studies to reduce the increasing rate of overweight and obesity.

## Introduction

Being overweight is a condition where the body has excessive food deposits than necessary. On the contrary, obesity is when excessive food deposits can hamper one’s health [1]. WHO defines overweight as a BMI greater or equal to 25, and obesity is a BMI greater than or equal to 30 for any adult. In 2022, 2.5 billion adults aged 18 years and older were overweight, including over 890 million adults who were living with obesity [1]. Similarly, in Bangladesh, the proportion of women who are overweight or obese increased from 3% in 1997 to 32% in 2018 [2]. Thus, the country needs to explore the determinants of this increasing trend. Studies around the world found that age group, education, occupation, employment status, monthly income, white-collar job, socioeconomic status, diet, physical activity, marital status, residential area, and sedentary behaviors are responsible for overweight and obesity among individuals [3–10].

Several studies have indicated that watching television works as a sedentary behavior to increase overweight and obesity [3–5,11]. Watching television decreases PA and increases levels of sitting/lying, junk food consumption. Moreover, Individuals’ risk of overweight and obesity differs with their occupation types [4,5,12]. White-collar job is associated with increasing obesity and overweight [7,13–15]. Studies have also indicated that blue-collar workers report higher levels of physical activity than white-collar workers [9,15]. On the contrary, white-collar workers reported the highest stress and procrastination levels in their jobs and lives, which caused more overweight and obesity [8,13]. White-collar workers showed high procrastination levels, which is positively associated with BMI. Moreover, jobseekers were less likely to be overweight than never-unemployed participants [16–18].

Globally, women (40%) are at more risk of being overweight and obese than men (35%) [19]. The gender discrepancy in overweight and obesity is aggravated among women in developing countries [20]. Many biological, contextual, and socio-cultural factors are responsible for this situation [19–21]. Obesity has many consequences for women, like fertility problems, gestational diabetes, gestational hypertension, preeclampsia, obstructive sleep apnea, lifetime hormonal changes, higher risk of cardiovascular diseases, type 2 diabetes, and breast cancer [19]. For this reason, it is necessary to determine the factors responsible for increasing overweight and obesity among women.

Obesity and overweight have become a crucial topic of interest for study in Bangladesh. Studies have focused on determinants, prevalence, and factors associated with overweight and obesity [10,22,23]. Only one study has focused on TV watching as a determinant of overweight and obesity in the country [23]. However, only one study focused on employment status as a determinant of overweight and obesity [24]. None of the studies has focused on the combined effect of sedentary behavior, television watching, unemployment, and white-collar jobs as determinants of overweight and obesity in the country. In this situation, our study aims to explore the effect of watching television, unemployment, and white-collar jobs on the overweight and obesity of reproductive-aged women (15-49) in Bangladesh.

## Data and Methods

### Source of Data and Inclusion Criteria

We applied the Bangladesh Demographic and Health Survey (BDHS) data*, 2017-18* [25]. The sample for the BDHS is nationally representative, and a detailed methodology will be found elsewhere in the report [25]. The BDHS follows two-stage stratified sampling with a response rate of 98.8%. **Figure 1** shows the sample selection process of this study. In the first stage, 675 (250 in urban and 425 in rural) enumeration areas (EAs) were selected with probability proportional to size. The survey was successfully carried out in 672 clusters after eliminating three clusters (one urban and two rural) that were eroded entirely by floodwater. Among the 20,160 households selected, interviews were completed in 19,457 occupied households. Among the 20,376 ever-married women aged 15-49 eligible for interviews, 20,127 were interviewed. The principal reason for non-response among women was their absence from home despite repeated visits. Response rates did not vary notably by urban-rural residence. The complete information on height or weight was for 19,786 women. Around 1635 women were excluded from the analysis due to current pregnancy, postpartum period (given birth two months before the survey), and flagged cases (BMI below 12.0 or above 60.0 are flagged). The final sample size for data analysis was 18151 ever-married women aged 15-49.

**Figure 1:**
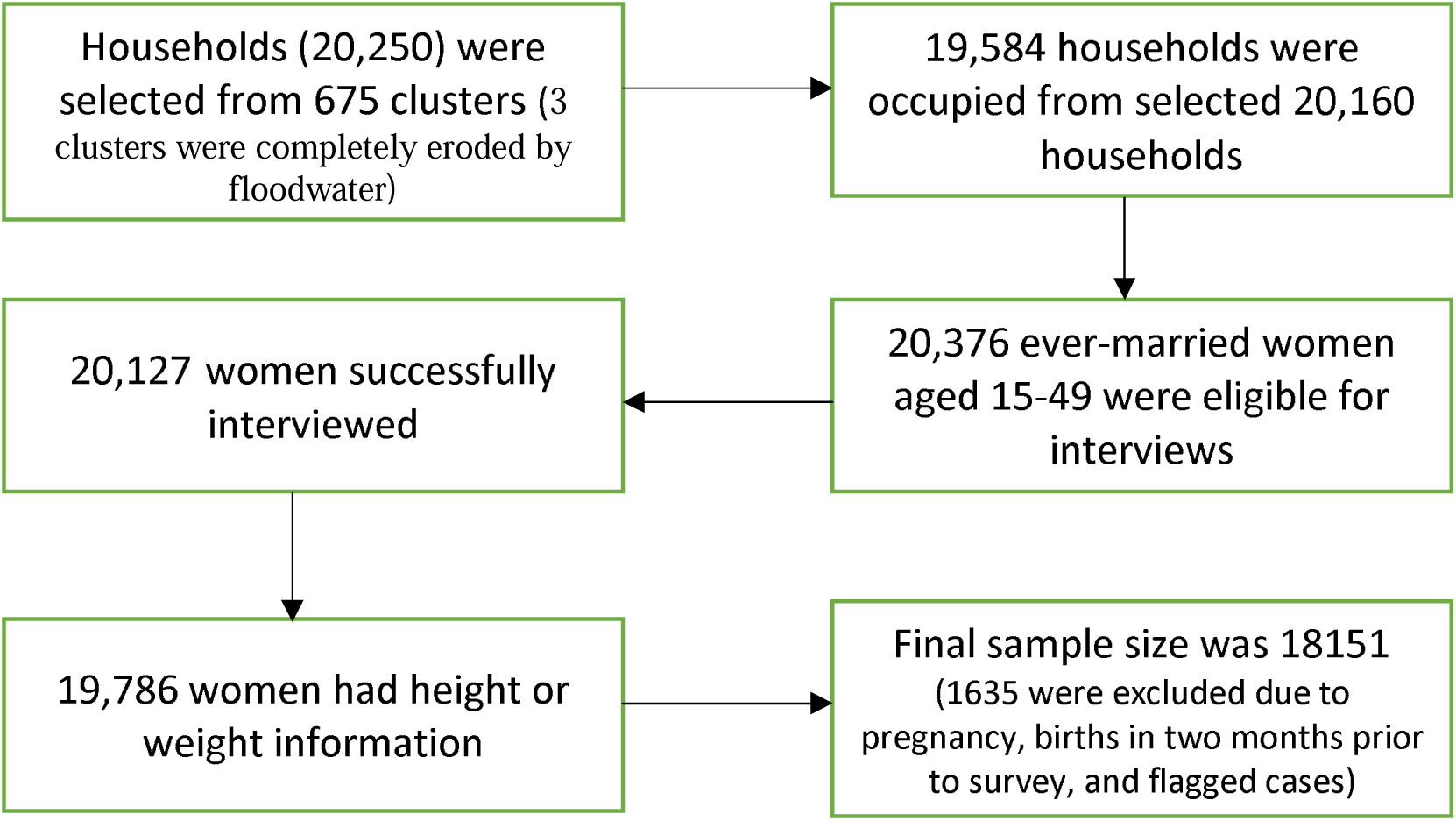
Process of Sample Selection for the Study

**Figure 2:**
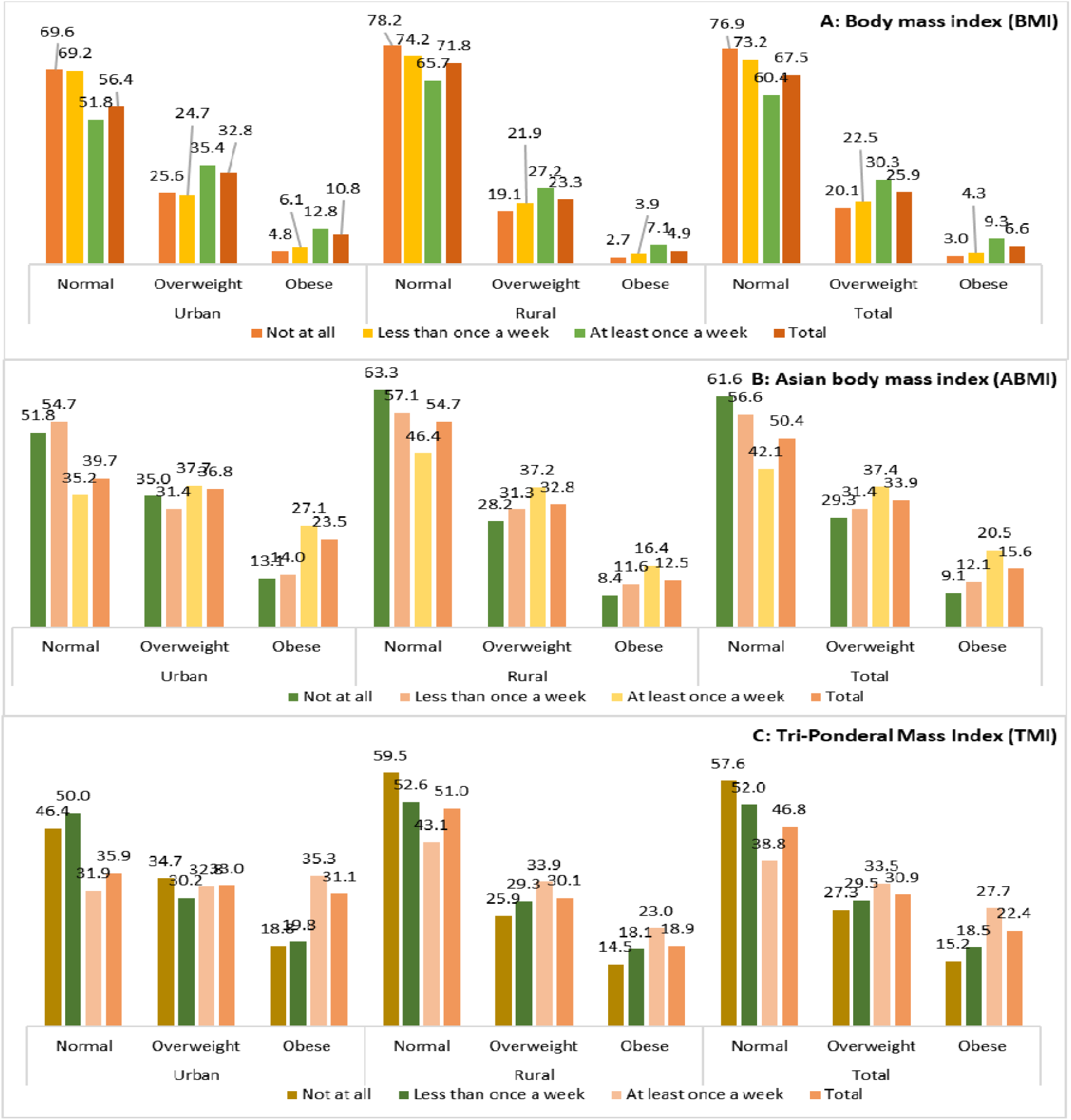
Nutritional status of ever-married women aged 15-49 in Bangladesh by frequency of watching television (%), 2017-18

**Figure 3:**
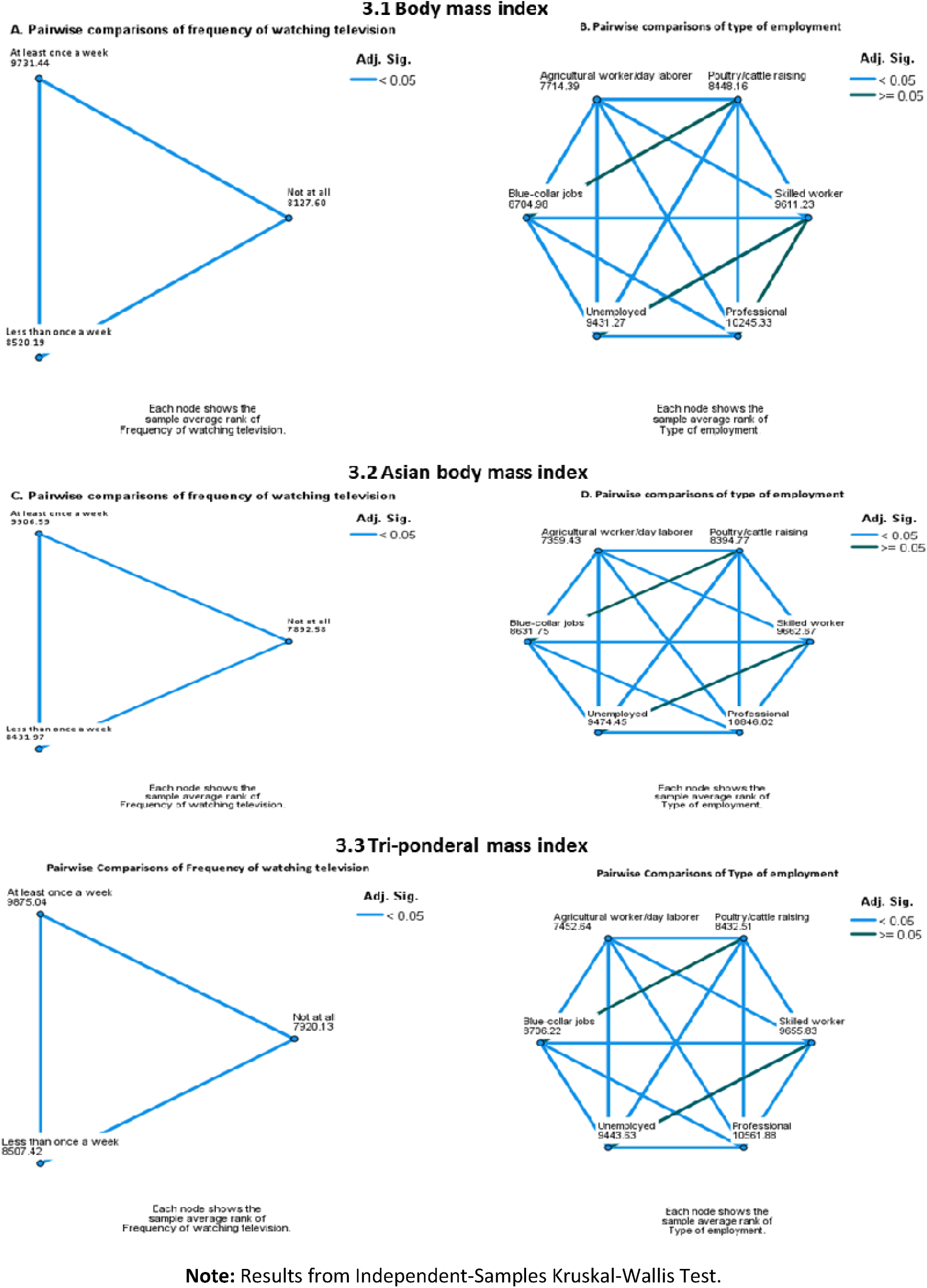
Pairwise comparison of nutritional status of ever-married women aged 15-49 in Bangladesh by frequency of watching television and types of employment, 2017-18

### Measures

#### Dependent variable

The dependent variable of the study was the nutritional status of women. We utilized three different measures: body mass index (BMI) of international standard, Asian body mass index (ABMI), and tri-ponderal mass index (TMI) as the outcome variable of the study. BMI was categorized as underweight/normal (≤24.99), overweight (25.0-29.99), and obese (≥30.0); whereas ABMI as underweight/normal (≤22.99), overweight (23.0-27.49), and obese (≥27.5). We used the categories of TMI as follows: underweight/normal (≤14.99), overweight (15.0-17.49), and obese (≥17.5).

### Independent variables

The frequency of watching television (TV) and types of employment were the two predictors of overweight and obesity in this study. The precise durations (hours or minutes/day) of watching TV were not available in the dataset, therefore, the following frequencies per week were used: (a) not watching TV at all, (b) watched less than once, and (c) watched at least once. We categorized types of occupation based on the nature of work as (a) not working (unemployed), (b) agricultural worker/ fisherman/rickshaw driver/ day laborer/ domestic servant, (c) poultry raising/ cattle raising/ farming, (d) home-based manufacturing/ blue-collar jobs, (e) skilled manual worker/small business, and (f) professional/ technical/ managerial/ big business/landowner. A white-collar worker was a person who performed professional, managerial, desk, or administrative work. In this study, skilled manual workers/small business and professional/ technical/ managerial/ big business/ landowner were considered white-collar jobs.

### Control variables

We selected covariates based on the existing literature [26–30] and the availability of variables in the dataset [25]. We included age, parity, marital status, religion, administrative division, place of residence, education, empowerment, household wealth index, and household size as covariates. We categorized the current age of women into 15-24, 25-34, and 35-49 years, which was also used in other studies [26,27,30]. Religion was categorized as Muslim and non-Muslim (Hindu, Buddhist, or Christian) due to most Muslims in Bangladesh.

Education was categorized as no education, primary incomplete, primary complete, secondary incomplete, and secondary complete and higher. The wealth index, which is used to assess the socioeconomic status of the household, was constructed from data on household possessions using the principal component analysis and divided into three groups (poor: bottom 40%, middle: next 40%, and rich: top 20%) based on overall asset ownership. We measured women’s participation in household decision-making as a proxy of women’s empowerment using four items: *the person who usually decides on the respondent’s health care* , *the person who usually determines on large household purchases* , *the person who usually decides on visits to family or relatives*, and *person who usually decides what to do with money husband earns* . The Cronbach’s alpha value was 0.81, indicating good internal consistency. We categorized empowerment as high if all the decisions were made by the respondents alone, moderate if all the decisions were made jointly with the husband or others, and low if husbands or others made the decision. We also measured community socioeconomic status by averaging individual education and household wealth in the PSU.

### Statistical Analysis

We performed the Wilcoxon-Mann-Whitney test, the Kruskal-Wallis test, and the Multilevel Ordered Logistic Regression Analysis as the outcome variable at the ordinal level. DHS data has a hierarchical structure with different levels: individuals nested within communities, and individuals within a cluster might be more similar than individuals in the rest of the country. It implies that we need to consider the variability between clusters. We used multilevel (mixed-effects) multiple-ordered logistic regression analyses to identify the effect of watching TV, unemployment, and white-collar jobs on overweight and obesity in Bangladesh. We used four models (null model, model using only frequency of watching TV, model using only the occupation types, and final model) to estimate the individual and community-level factors and random intercept of between-cluster variation.

The null model was to test the random effect of between-cluster variability without any independent variable. The intraclass correlation coefficient (ICC) was derived from the between-cluster and within-cluster variability. The only independent variable models examined the effects of exposure on nutritional status. Besides, the ICC was observed if there was a decline in the between-cluster variability after adding predictors to the null model. Finally, both predictors and other covariates and cluster variables were concurrently entered into one model (final) to reveal their net fixed and random effects. We used the IR (Individual Women’s Recode) file for multilevel analysis in Stata 17.

We used the variance inflator factor (VIF) to examine the instability of the effect size of predictors as the result of multicollinearity among the independent variables. The multiple analyses excluded the community-level education and wealth (VIF >10). The random effects were reported as the ICC, the percentage variance explained by the higher level (community-level variables). The proportional change in community variance (PCV) expresses the change in the community-level variance between the null model (Model 1) and the consecutive models (presented final model only). The estimated effects are presented as Adjusted Odds Ratio (aOR) with their standard errors (SE).

## Results

### Characteristics of the participants

The characteristics of the study participants are presented in **Table 1** . About 36% of the respondents did not watch TV at all, whereas 55% watched at least once a week. Almost half of the women were unemployed, and 32.9% were involved in poultry raising/ cattle raising/ farming. Most participants (40.2%) were between the ages of 35 and 49 years. Around 57% were female and 80% were currently married. About 26% of the respondents had no institutional education, and 60.5% were employed. Around 73% of the respondents were from rural areas, and 23.9% were from Dhaka.

**Table 1:** Characteristics of the participants (%), 2017-18.

| Background characteristics | Sample, % (n) | Background characteristics | Sample, % (n) |
| --- | --- | --- | --- |
| **Frequency of watching television** |  | **Highest educational level** |  |
| Not at all | 35.7 (6486) | No education | 17.7 (3210) |
| Less than once a week | 9.3 (1683) | Primary | 31.7 (5759) |
| At least once a week | 55.0 (9982) | Secondary | 38.8 (7037) |
| **Type of occupation** |  | Higher | 11.8 (2145) |
| Agricultural worker <sup>a</sup> | 3.9 (713) | **Women empowerment** |  |
| Poultry raising/cattle raising/farming | 32.9 (5970) | Low | 16.9 (3063) |
| Home-based manufacturing/ blue-collar jobs <sup>b</sup> | 6.0 (1091) | Moderate | 77.4 (14050) |
| Professional <sup>c</sup> | 1.8 (323) | High | 5.7 (1037) |
| Skilled manual worker/small business <sup>d</sup> | 7.1 (1285) | **Parity** |  |
| Not working | 48.3 (8769) | 0 | 8.9 (1610) |
| **Age of the Respondent** |  | 1 | 21.5 (3909) |
| 15-24 | 24.9 (4526) | 2 | 31.7 (5752) |
| 25-34 | 34.9 (6329) | 3 | 21.2 (3852) |
| 35-49 | 40.2 (7297) | 4+ | 16.7 (3030) |
| **Marital status** |  | **Household size** |  |
| Currently in union | 93.9 (17036) | 1-4 | 45.5 (8263) |
| Formerly in union | 6.1 (1115) | 5-6 | 32.6 (5926) |
| **Religion** |  | 7+ | 21.8 (3963) |
| Islam | 90.4 (16418) | **Household wealth index** |  |
| Others | 9.6 (1734) | Poor | 38.4 (6963) |
| **Division** |  | Middle | 41.1 (7465) |
| Barisal | 5.6 (1014) | Rich | 20.5 (3723) |
| Chittagong | 17.7 (3221) | **Type of area** |  |
| Dhaka | 25.1 (4553) | Rural | 71.8 (13028) |
| Khulna | 11.9 (2153) | City Corporation | 7.1 (1294) |
| Mymensingh | 7.6 (1387) | Other urban | 21.1 (3829) |
| Rajshahi | 14.2 (2584) | **Total** | 100.0 (18151) |
| Rangpur | 12.1 (2196) |  |  |
| Sylhet | 5.8 (1045) |  |  |

**a=** Agricultural worker, Fisherman, Rickshaw driver, Brick breaking, Road building, Construction worker, Boatman, and Earth work, etc.; **b=**Home-based manufacturing, blue-collared jobs (Factory worker); **c=**Professional, Technical, Managerial, Big business, and Land owner; **d=** Skilled manual worker (Carpenter, Masson, Bus/taxi driver, Construction supervisor, Seamstresses/Tailor, Policeman, Armed services, Dai, Commun) and Small business **c** and **d** indicate white-collar jobs as they were related to professional, managerial, desk, less physical activity, or administrative work.

### Nutritional status by watching television and type of occupation

The prevalence of overweight among women who watched television at least once a week is higher than among women who did not watch television at all (30.3% vs 20.1%). The prevalence was higher among urban women than rural women (32.8% vs 23.3%). Again, the prevalence was higher among women who watched television once a week than women who did not, both in urban (35.4% vs 25.6%) and rural (27.2% vs 19.1%).

The prevalence of obesity showed a similar pattern. Women who watched television once a week had a higher prevalence of obesity (9.3%) than women who did not watch television at all (3.0%). Urban women were more obese than rural women (10.8% vs 4.9%). However, women who watched television were more likely to be obese both in urban and rural areas. Furthermore, the prevalence of overweight and obese differs substantially from the body mass index (BMI) of international standards, Asian body mass index (ABMI), and tri-ponderal mass index (TMI). Nevertheless, the pattern was the same on every scale.

In terms of overweight, women who watched television at least once a week were more likely to be overweight than women who did not watch television at all in both ABMI (37.4% vs 29.3%) and TMI (33.5% vs 27.3%). The pattern was consistent for obesity, too. The value for ABMI was (20.5% vs 9.1%) and for TMI (27.7% vs 15.2%). Urban women were more likely to be overweight and obese by both standards.

Women in agricultural work were less likely to become overweight (16.3% vs 28.7%) and obese (2.9% vs 8.2%) than those who were not working (**Table 2**). The white-collar job (professional) was working as an exception here. Women working in professional positions were most likely to be overweight and obese (35.4% and 10.9%). The pattern for professional positions was the same for ABMI (41% and 27.3%) and TMI (38.1% and 31.9%). The women engaged in agricultural work were less likely to be overweight (25.4% vs 35.0%) and obese (8% vs 18.8%) than women who were not working, according to ABMI. Similarly, women engaged in agricultural work were less likely to be overweight (24.8% vs 30.6%) and obese (13.6% vs 26.2%) than women who were not working by TMI. Urban women were more likely to be overweight and obese than rural women in every kind of job by BMI and PMI.

**Table 2:** Nutritional status of ever-married women aged 15-49 in Bangladesh by occupation types (%). 2017-18.

| Occupation | Urban |  |  | Rural |  |  | Total |  |  |
| --- | --- | --- | --- | --- | --- | --- | --- | --- | --- |
|  | Normal | Overweight | Obese | Normal | Overweight | Obese | Normal | Overweight | Obese |
| **Body Mass Index (BMI)** |  |  |  |  |  |  |  |  |  |
| Agricultural worker <sup>a</sup> | 75.9 | 20.3 | 3.8 | 84.2 | 13.5 | 2.4 | 80.8 | 16.3 | 2.9 |
| Poultry raising/cattle raising/farming | 66.7 | 27.7 | 5.6 | 74.4 | 21.6 | 4.0 | 73.6 | 22.2 | 4.2 |
| Home-based manufacturing/ blue-collar jobs <sup>b</sup> | 66.0 | 27.4 | 6.6 | 76.9 | 18.7 | 4.4 | 71.1 | 23.4 | 5.5 |
| Professional <sup>c</sup> | 47.0 | 42.1 | 11.0 | 60.4 | 28.9 | 10.7 | 53.7 | 35.4 | 10.9 |
| Skilled manual worker/small business <sup>d</sup> | 56.1 | 31.9 | 11.9 | 66.2 | 27.2 | 6.6 | 61.9 | 29.3 | 8.9 |
| Not working | 50.8 | 35.9 | 13.3 | 69.1 | 25.2 | 5.6 | 63.1 | 28.7 | 8.2 |
| **Asian Body Mass Index (ABMI)** |  |  |  |  |  |  |  |  |  |
| Agricultural worker <sup>a</sup> | 59.9 | 28.4 | 11.8 | 71.2 | 23.4 | 5.4 | 66.6 | 25.4 | 8.0 |
| Poultry raising/cattle raising/farming | 47.7 | 37.0 | 15.3 | 57.5 | 31.8 | 10.7 | 56.4 | 32.4 | 11.2 |
| Home-based manufacturing/ blue-collar jobs <sup>b</sup> | 49.9 | 35.5 | 14.6 | 60.0 | 27.9 | 12.2 | 54.6 | 32.0 | 13.5 |
| Professional <sup>c</sup> | 28.8 | 37.4 | 33.7 | 34.6 | 44.7 | 20.8 | 31.7 | 41.0 | 27.3 |
| Skilled manual worker/small business <sup>d</sup> | 35.4 | 41.5 | 23.1 | 50.5 | 36.1 | 13.4 | 44.0 | 38.4 | 17.6 |
| Not working | 35.3 | 36.9 | 27.8 | 51.5 | 34.1 | 14.4 | 46.2 | 35.0 | 18.8 |
| **Tri-Ponderal Mass Index (TMI)** |  |  |  |  |  |  |  |  |  |
| Agricultural worker <sup>a</sup> | 53.1 | 28.6 | 18.3 | 67.2 | 22.4 | 10.4 | 61.6 | 24.8 | 13.6 |
| Poultry raising/cattle raising/farming | 42.6 | 35.0 | 22.4 | 53.1 | 30.2 | 16.6 | 52.0 | 30.7 | 17.2 |
| Home-based manufacturing/ blue-collar jobs <sup>b</sup> | 44.6 | 34.1 | 21.4 | 54.6 | 28.9 | 16.5 | 49.1 | 31.7 | 19.2 |
| Professional <sup>c</sup> | 27.9 | 33.9 | 38.2 | 32.3 | 42.4 | 25.3 | 30.0 | 38.1 | 31.9 |
| Skilled manual worker/small business <sup>d</sup> | 31.9 | 36.6 | 31.5 | 47.2 | 32.4 | 20.4 | 40.6 | 34.2 | 25.1 |
| Not working | 32.1 | 32.1 | 35.8 | 48.6 | 29.9 | 21.5 | 43.2 | 30.6 | 26.2 |

**a=** Agricultural worker, Fisherman, Rickshaw driver, Brick breaking, Road building, Construction worker, Boatman, and Earth work etc.; **b=**Home-based manufacturing, blue-collared jobs (Factory worker); **c=**Professional, Technical, Managerial, Big business, and Land owner; **d=** Skilled manual worker (Carpenter, Masson, Bus/taxi driver, Construction supervisor, Seamstresses/Tailor, Policeman, Armed services, Dai, Commun) and Small business. **c** and **d** indicate white- collar jobs.

### Effect of watching television, unemployment, and white-collar jobs on nutritional status

Women who watched television less than once a week were not significantly associated with BMI (p=0.653), ABMI (p=0.388), and TMI (p=0.221) compared to those who did not watch at all (**Table 3**). The results showed a significant association between women who watched television at least once a week and their BMI (36% higher than those who did not watch at all), ABMI (42% higher), and TMI (39% higher). The association between professional work (white-collar job) with BMI, ABMI, and TMI was statistically significant. Women in professional work had higher BMI (62% than agricultural workers/ day laborers), ABMI (81% higher), and TMI (67% higher). In the same fashion, the association between women who were not working (unemployment) and BMI, ABMI, and TMI is statistically significant. Unemployed women had 106 times higher BMI than agriculture workers/ day laborers and had 91% higher ABMI and 86% higher TMI.

**Table 3:** The effect of watching TV, unemployment, and white-collar jobs on overweight and obesity among ever-married women aged 15-49 in Bangladesh using multilevel ordered logistic regression analysis.

|  | Body Mass Index,<br>aOR (SE) | Asian Body Mass Index,<br>aOR (SE) | Tri-Ponderal Mass Index,<br>aOR (SE) |
| --- | --- | --- | --- |
| **Watching television** |  |  |  |
| Not at all | [RC] | [RC] | [RC] |
| Less than once a week | 1.04 [0.08] | 1.06 [0.07] | 1.08 [0.07] |
| *P-value* | 0.653 | 0.388 | 0.221 |
| At least once a week | 1.36 [0.07] | 1.42 [0.06] | 1.39 [0.06] |
| *P-value* | <0.001 | <0.001 | <0.001 |
| **Occupation** |  |  |  |
| Agricultural worker/day laborer <sup>a</sup> | [RC] | [RC] | [RC] |
| Poultry raising/cattle raising <sup>b</sup> | 1.53 [0.19] | 1.51 [0.17] | 1.49 [0.17] |
| *P-value* | <0.001 | <0.001 | <0.001 |
| Home-based manufacturing <sup>c</sup> | 1.37 [0.20] | 1.31 [0.18] | 1.31 [0.16] |
| *P-value* | 0.034 | 0.046 | 0.028 |
| Professional <sup>d</sup> | 1.62 [0.29] | 1.81 [0.31] | 1.67 [0.27] |
| *P-value* | 0.007 | 0.001 | 0.002 |
| Skilled worker/small business <sup>e</sup> | 1.99 [0.26] | 1.81 [0.22] | 1.80 [0.21] |
| *P-value* | <0.001 | <0.001 | <0.001 |
| Not working | 2.06 [0.25] | 1.91 [0.21] | 1.86 [0.19] |
| *P-value* | <0.001 | <0.001 | <0.001 |
| **Cut1: Null model** | 0.79 [0.03] | 0.02 [0.03] | -0.14 [0.02] |
| **Cut1: Final model** | 3.34 [0.17] | 2.49 [0.15] | 2.17 [0.14] |
| **Cut2: Null model** | 2.81 [0.04] | 1.80 [0.03] | 1.33 [0.03] |
| **Cut2: Final model** | 5.49 [0.17] | 4.42 [0.15] | 3.74 [0.15] |

| Model Summary |  |  |  |
| --- | --- | --- | --- |
| Variance: Null model | 0.39 [0.03] | 0.37 [0.03] | 0.32 [0.03] |
| Variance: Final model | 0.06 [0.01] | 0.05 [0.01] | 0.05 [0.01] |
| ICC: Null model | 0.105 | 0.100 | 0.099 |
| ICC: Final model | 0.018 | 0.016 | 0.014 |
| PCV: Final model | 84.6 | 85.2 | 85.4 |

**a**=Agricultural worker/rickshaw driver/day laborer/domestic servant; **b**=Poultry raising/cattle raising/farming; **c**=Home-based manufacturing/ blue-collared jobs (non-agricultural worker); **d**=Professional/ technical/ managerial/ big business/ landowner; **e**= Skilled manual worker/ small business. **d** and **e** indicate white-collar jobs.
aOR=Adjusted Odds Ratio: adjusted for age, marital status, parity, household size, religion, education, women empowerment, household wealth index, place of residence, and administrative divisions.
SE=Standard error.

## Discussion

This study attempted to show whether television watching, unemployment, and white-collar jobs are working as a sedentary behavior to increase overweight and obesity among reproductive-aged women. Based on nationally representative cross-sectional data, we observed an increasing trend in overweight and obesity among reproductive-aged women in Bangladesh. The prevalence of overweight and obesity was higher among urban women than rural. It is a universal pattern observed for all occupation groups of women, whether in agricultural work or women with professional jobs. Past studies of the country also showed the same pattern [31,32]. Urban women who are not working are more likely to be overweight and obese than their rural counterparts. This pattern is consistent with TV watching, too. Past studies of many countries indicated that the difference between urban and rural is a consequence of the high exposure of urban women to technology, diverse TV channels, urbanization, and more unhealthy food [33,34].

Women who have watched television at least once a week are more likely to have a BMI ≥25 and become overweight and obese than those who didn’t watch TV at all. Studies around the world have found a significant relationship between overweight and obesity and the frequency of watching television. One survey in India reported that women who watched television had the highest score of ABMI [3]. Again, a study in Ethiopia found that watching television at least once a week was significantly associated with both overweight and obesity [6]. Furthermore, studies of Nepal, Ghana, Myanmar, and Peru also found the same result using BMI and ABMI as indicators for measuring overweight and obesity [35–38].

This same pattern of BMI is discernible among unemployed women compared to women in agricultural work. One study in Bangladesh indicated that unemployed women had the highest risk of being overweight and obese compared to their counterparts [24]. Studies have found perplexing results on this topic. A study in Brazil, Russia, India, China, and South Africa found an association between unemployment and overweight and obesity. However, in India, employment status affected overweight and obesity, while the relationship was inverse for China [16]. Most of the studies of other countries and past research of our country have focused on how obesity affects unemployment and job loss [24,39,40]. Thus, our study has demonstrated a unique pathway in the relationship between employment status and overweight and obesity.

Finally, our study found that professional and white-collar job women have the highest risk of being overweight and obese. Studies have reported contradictory results in this context. While some studies reported an association between blue-collar jobs and lower levels of overweight and obesity, other studies reported blue-collar jobs to have a higher risk of overweight or obesity compared to white-collar jobs [8,9,17].

### Strength and limitations

This study is the first attempt in Bangladesh to identify whether unemployment and occupation status are sedentary behaviors in reproductive-aged women. Another study has demonstrated the effect of TV watching on overweight and obesity in Bangladesh, according to BDHS-2014. However, this study has revised its findings, used three international measurement standards, and presented its relationship with sedentary behavior, which gives strong evidence against our conclusion. Not only that, but this also provides knowledge about the degree and extent of variations among different standards. This study has one crucial limitation. It has presented results only for reproductive-aged women. Because readily available data is not available for men and other age group. Future research investigations can focus on this aspect.

## Conclusion

Worldwide, an increasing number of people are becoming overweight and obese. Bangladesh is following the same trend. In this regard, studies need to investigate the relationship between different behavior and its effect on overweight and obesity in Bangladesh. Policymakers need to guide their focus on making routine data available for all age groups. Thus, more studies are needed to guide this topic. Awareness programs need to be shaped to encourage people to avoid behavior like junk food and beverages and increase their physical activity. An occupational structure needs to be created, and an environment that promotes healthy behavior for women needs to be made. Not only that, but appropriate equipment and a gender-sensitive structure also need to be guided. Urban women are more prone to overweight and obesity. Special programs and social services like gyms and parks must be guided for urban women. Facilities used by reproductive-aged women, like hospitals and childcare centers, need to take a leading role in spreading awareness about it. On top of all that, women need to prioritize their health and take actions that benefit them.

## Author Approval

All authors have seen and approved the manuscript.

## Declarations Ethics approval

The National Institute of Population Research and Training (NIPORT) of the Ministry of Health and Family Welfare conducted the 2017–18 BDHS. The survey was implemented by a Bangladeshi firm named Mitra and Associates of Bangladesh. At the same time, the ICF International of USA provided technical assistance as part of its international Demographic and Health Surveys Program. If the respondent provided their verbal consent in response to being read out an informed consent statement by the interviewer, only then was an interview conducted. The ethical approval for the survey was taken by the NIPORT from the Bangladesh Medical Research Council (BMRC).

## Patient consent for publication

Not applicable.

## Data availability statement

The dataset (BDHS 2017-18) used in this study is publicly available on the DHS website The DHS Program - Available Datasets.

## Acknowledgment

We acknowledge the Measure DHS (Demographic and Health Surveys) Data Archive (URL: https://dhsprogram.com/data), ICF International, the USA for granting access to the Bangladesh Demographic and Health Survey data.

